# Pattern and association of immunoglobulin G antibodies to AMA1, GLURP, and MSP3 with protection from malaria in a cohort of Cameroonian children living in Mutengene: Anaemia a possible collateral damage?

**DOI:** 10.1101/2025.01.04.24319738

**Authors:** Clarisse Njua-Yafi, Eric A. Achidi, Judith K. Anchang-Kimbi, Tobias O. Apinjoh, Regina N. Mugri, Hanesh F. Chi, Rolland B. Tata, Charles Njumkeng, Daniel Dodoo

## Abstract

Understanding the characteristics of naturally acquired immunity in different epidemiologic settings is essential for vaccine development and testing. The relationship between antibodies against four malaria vaccine candidate antigens and protection from malaria in a cohort of Cameroonian children was assessed. Immunoglobulin (Ig)G and IgG subclasses against recombinant *Plasmodium falciparum* apical membrane antigen 1 (AMA1), glutamate rich protein (GLURP) R0, GLURP R2 and merozoite surface protein 3 (MSP3) in the plasma of 357 Cameroonian children were measured by sandwich ELISA at three time points (baseline, 6 months and 12 months) during which time participants were monitored for malaria.Total IgG to all four antigens correlated positively with age (0.51≤ r ≤ 0.23, p < 0.001) at all three time points. Adjusting for age, total IgG, IgG1, IgG3, IgG2 (except for MSP3 IgG2) antibody levels to all four antigens were associated with protection against malaria parasitaemia at baseline. GLURP R0 IgG (F = 35.7, p < 0.001), GLURP R2 IgG (F = 16.5, p < 0.001), AMA1-3D7 IgG2 (F = 10.8, p < 0.001) and AMA1-3D7 IgG3 (F = 4.01, p = 0.019) decreased with a corresponding decrease in malaria cases (χ^2^ = 10.4, p = 0.034) across the three time points, contrary to the increase observed in MSP3 IgG (F = 8.9, p < 0.001) and MSP3 IgG2 (F = 44.2, p < 0.001). Increased levels of AMA1-3D7 IgG [OR = 4.13, 95% CI (1.09 – 15.65), p = 0.037] and MSP3 IgG1 [OR = 8.16, 95% CI (1.06 – 62.64), p = 0.044] were associated with susceptibility to anaemia after controlling for age and parasitaemia.Total IgG, cytophilic subclasses and IgG2 to all the antigens (except MSP3 IgG2) were associated with malaria protection while MSP3 IgG seemed to persist longer. The relationship between malaria specific antibodies and anaemia warrants further studies.

## Introduction

An increase in malaria control and elimination strategies has resulted in many endemic areas transitioning from high-to-low-to-no malaria transmission. The Mount Cameroon region of South Western Cameroon which was previously hyper-endemic for malaria [1, 2] is currently meso-endemic [3,4,5]. Despite advances in the fight against malaria, it still remains a major public health problem especially in Sub-Saharan Africa which accounted for about 95% of the global malaria cases and deaths in 2021 and the death of nearly half a million African children under the age of five years [6]. The acquisition of immunity to malaria in populations is evident by the declining incidence of uncomplicated and severe malaria with increasing age. Reductions in transmission will have an impact on the development of naturally acquired immunity to malaria which develops after repeated exposure to *Plasmodium* spp. [7].

Antibodies constitute an important component of naturally acquired as well as blood-stage immunity, as demonstrated by experimental animal models and the passive transfer of IgG from malaria immune African adults to partially immune African or Thai children which rapidly reduced parasitaemia and fever [8, 9, 10]. Naturally acquired malaria-specific antibodies, particularly cytophilic IgG (IgG1 and IgG3) have frequently been associated with protection against *Plasmodium falciparum* malaria [1, 11, 12, 13, 14]. In other studies, IgG2 antibodies to certain *P. falciparum* antigens in the presence of a receptor polymorphism that favours IgG2 binding have been associated with protection against malaria [15, 16].

The pre-erythrocytic RTS,S/AS01 malaria vaccine is the first and only malaria vaccine recommended by WHO for use to prevent *P. falciparum* malaria in children living in regions with moderate-to-high malaria transmission [17]. This came after pilot implementations in Ghana, Kenya and Malawi which began in 2019 showed that the vaccine substantially reduced deadly severe malaria, decreased hospitalizations and also resulted in a drop in child deaths, according to WHO [18]. Two other pre-erythrocytic vaccine candidates are approaching late-stage clinical evaluation: the R21/MatrixM vaccine and the attenuated whole sporozoite vaccine PfSPZ [17]. Having two or more vaccines improves vaccine security thereby underscoring the need for sustained efforts towards more efficacious malaria vaccines. Since the major malaria pathology is associated with erythrocytic parasite replication, a protein that elicits invasion-blocking antibodies would represent an obvious complement to the sporozoite antigen *Pf*CSP [19]. Several blood-stage antigens including the apical membrane antigen 1 (AMA1), merozoite surface protein (MSP) 1, MSP2, MSP3, glutamate rich protein (GLURP), serine repeat antigen (SERA5) and the erythrocyte-binding antigen 175 (EBA 175) have been clinically evaluated as vaccines [20, 21, 22, 23, 24, 25, 26, 27, 28]. Unfortunately, phase II trials of some advanced blood-stage candidates, (AMA1 and MSP1) did not demonstrate efficacy in African children [29, 30]. The *P. falciparum* MSP3 and glutamate-rich protein (GLURP) fusion protein (GMZ2) made good progress in clinical trials demonstrating an efficacy of 14% in children aged less than 5 years in a phase IIb trial [31, 32]. The most promising blood-stage vaccine candidate so far is the *P. falciparum* reticulocyte-binding protein homologue 5, PfRH5 [33]. It is widely accepted that a combination vaccine for malaria (especially multi-stage) is likely to be more effective than vaccines based on a single antigen, however, the design of a multi-component vaccine requires careful assessment to avoid possible immune interference. Understanding the characteristics and components of naturally acquired immunity to different antigens in different epidemiologic settings is essential for vaccine development as well as for testing vaccine candidates for immunogenicity and protective efficacy [34]. Thus, we assessed the levels of total IgG and IgG subclasses to the recombinant antigens AMA1, GLURP R0, GLURP R2 and MSP in relation to asymptomatic and symptomatic *P. falciparum* infection in a longitudinal cohort study of Cameroonian children residing in Mutengene.

## Materials and methods

### Study area

The study was conducted in Mutengene, a semi-urban community located in the Mount Cameroon region, South West Region of Cameroon between February 2011 and March 2012. The study community comprised ten residential quarters spanning the entire Mutengene. The community has a Medical Integrated Health Centre which is the only government owned institution that offers affordable health services to the community. Consenting volunteer participants were sampled either at the Mutengene Medical Integrated Health Centre or at mobile temporary sampling sites within the community. The Mutengene health area has an equatorial climate that consists of a short dry season from late November to February and a long rainy season from March to November with maximum rainfall between June and August interspersed by early moderate (March – May) and late moderate (September – November) rains [35]. Mutengene was characterized by mean temperatures of 25.1°C and a mean relative humidity of 83.1%, according to the CDC (Cameroon Development Corporation) weather records then [36]. The Mt. Cameroon region which was previously hyper-endemic for malaria [1, 2] with a very high entomological inoculation rate (EIR) reaching an annual average of 1077.1 infectious bites/person/year at Esuke Camp in Mutengene [37], is currently meso-endemic [5]. Malaria transmission is more intense during the rainy season with peak transmission during the heavy rains. The main malaria vectors are *Anopheles gambiae, An. funetus and An. hancocki. P. falciparum* is the predominant malaria parasite species and accounts for up to 96.0% malaria infections in the study area [38].

### Study population and sampling procedure

This was a community-based one year (February 2011 to March 2012) longitudinal cohort study of children aged 6 months to 10 years from randomly selected households in the community. After randomly selecting 10 (50 %) of the 20 quarters in the study community, households within the selected quarters also went through systematic random selection. All the households in the study community were identified and numbered such that each household had an equal probability of being selected using systematic random sampling. An interval of 3 – 5 households with children less than 10 years old depending on the population density of each quarter was used for the selection of eligible households. When selection fell on a business structure or household without children under 10 years old or lack of consent from family head, an eligible household was selected ± 2 households around the concerned household. After voluntary informed consent was obtained, participants were recruited from households (at most two per household giving preference to younger children) resident in the community. The incidence of malaria was determined by both active case detection (biweekly home visits) and passive case detection through self-referral to the Mutengene Health Centre (or other health establishments in the community). During these home visits the health status of the children was assessed using a structured morbidity questionnaire and identified febrile children were brought to the health centre for clinical examination, diagnosis and treatment. Finger prick blood samples were collected from all febrile/sick children at the health centre for haemoglobin measurement and malaria parasitaemia (mp) determination.

Anthropometric measurements and axillary temperature of participants were recorded every three months from enrolment till one year and venous blood samples collected at enrolment, 6 months and 12 months for mp determination, full blood count, haemoglobin electrophoresis and *P. falciparum* antibody measurement by ELISA. An episode of malaria was defined as fever with an axillary temperature >37.5°C, the presence of malaria parasites in a thick blood film and one other sign/symptom of malaria (shaking chills, headache, muscle ache, vomiting or fatigue). Children presenting with acute malaria during the study period were referred to a physician where they were given artemeter-lumefantrine, a first line ACT recommended for use at that time in the treatment of uncomplicated malaria in Cameroon. All participants were de-wormed at enrolment and had access to free consultation, malaria tests and treatment throughout the study period. In addition to failure to obtain informed consent from a parent or guardian, children with sickle cell disease, any obvious congenital disorder or those who would be above 10 years of age by next birth day were excluded from the study.

### Haematological analysis

Full blood count was determined in venous blood using an automated haemoanalyser (Teco, USA). Anaemia was defined as any haemoglobin (Hb) level <11g/dl and further classified based on WHO guidelines as severe anaemia: Hb< 7g/dl, moderate anaemia: Hb 7 – 9.9g/dl, mild anaemia: Hb 10 – 10.9g/dl [39].

### Haemoglobin electrophoresis

The Hb genotypes were determined by cellulose acetate alkaline electrophoresis using lysed packed red cells as described by Cheesebrough [40]. Red blood cell samples and controls were lysed using the Hemolysate Reagent (W/O KCN) [HELENA Electrophoresis Systems, BIOSCIENCES]. The electrophoresis was performed in 1X Tris Glycine buffer in an electric field of 250 V for 25 - 30 minutes.

### Malaria parasitaemia determination

Thick and thin blood films prepared from blood samples following standard procedures were stained with 10 % Giemsa (Sigma, St Louis, USA). Malaria parasitaemia status, density and species were determined by light microscopy using an Olympus microscope (Olympus Optical Co., Ltd, Japan). Smears were reported as negative only after observing at least 100 high power fields. Using the white blood cell (WBC) count per μl of blood of each participant, the parasitaemia density/μl was estimated by counting the number of asexual parasites against a minimum of 200 leukocytes and calculated using the following formula:

Parasitaemia density (parasites/µl of blood) = (No of parasites × WBC count/µl)/(No of WBCs counted).

### Measurement of IgG and IgG subclasses to *P. falciparum* AMA1, MSP3 and GLURP (R0 & R2) antigens

IgG and IgG subclass (IgG1, IgG2, IgG3 and IgG4) antibodies to recombinant malaria antigens AMA1, MSP3, GLURP R0 and GLURP R2 were quantified by indirect ELISA using a modified version of the Afro Immuno Assay (AIA) ELISA protocol as described by Dodoo *et al*, [41]. Nunc-Immuno [(96-well) Thermo Fisher Scientific, USA] microtitre plates were coated (100 µl/well) with AMA1, MSP3 and GLURP R0 at 1µg/ml and GLURP R2 at 0.5µg/ml in PBS (pH 7.2) and kept overnight at 2 - 8°C. Plates were washed four times with PBS - 0.05%Tween 20 (wash buffer) using a microplate autowasher (BioTek Instruments, USA), padded dry on tissue paper and blocked (200µl/well) with PBS - 3% skim milk powder (Sigma-Aldrich) at room temperature for 1hr. Plasma samples and negative controls [eleven malaria non-exposed US volunteers (Martha Sedega’s group, US Naval Medical Research Centre)] were diluted 1/1000 for AMA1 and 1/200 for MSP3, GLURP R0 & R2 in serum dilution buffer (PBS, 1% milk powder, 0.02% Sodium Azide). To control for inter-assay and day-to-day variations in the standardized ELISA procedure, each assay (ELISA plate) included a calibration curve obtained by a 2-fold titration of a pool of hyper immune sera/plasma from malaria exposed Ghanaians known to be positive for the antibodies (IgG and IgG1 to IgG4) to the malaria antigens tested. In addition, each plate also included a negative control sample, a positive control sample and a buffer blank (serum dilution buffer) which served as internal controls to allow for detection of a failed assay run. Samples, standards and controls were added in duplicates (100µl/well) and the plates incubated for 2hrs at room temperature and then washed four times with PBS – 0.05%Tween 20.

The optimised dilutions (in PBS with 1% milk) for the detection antibodies used in the assays were; horse radish peroxidase (HRPO) conjugated goat anti-human IgG [Invitrogen – USA, (1/8000)], HRPO conjugated sheep anti-human IgG1 (1/3000), IgG2 (1/1000), IgG3 (1/3000) and IgG4 (1/2000) (The Binding Site, UK). The conjugates were added (100µl/well), plates incubated at room temperature for 1hr and washed four times with wash buffer. Bound secondary antibody was quantified by colouring (100µl/well) with ready to use TMB (3, 3’, 5, 5’-Tetramethylbenzidine) plus substrate (Kem-En-Tec Diagnosis, Denmark) and incubated in the dark for 10 to 15minutes. The reaction was stopped with 100µl/well of 0.2MH_2_SO_4_ and Optical densities (ODs) were read at 450 nm using the Biotek EL 808 ELISA plate reader (Biotek Instruments, USA). Optical density (OD) values for the test samples were converted into antibody units (AU) using the standard reference curves generated for each ELISA plate with a four parameter curve-fit Microsoft Excel-based application (ADAMSEL b040, Ed Remark^©^ 2009). Samples were re-tested if the coefficient of variation between duplicate absorbance values was higher than 15% and plates were also re-tested if the R-square value of the standard curve was less than 97%.

Antibodies to the antigens were measured at baseline (n = 358), 6 months (n = 289) and 12 months (n = 272). The reference standard curves used in transforming all optical density (OD) values to antibody units were optimized such that the same antibody unit of 3 was assigned to an OD of 0.3 in all antibody measurements irrespective of the antigen being tested.

### Statistical analysis

Data was entered into Microsoft Excel and analyzed using SPSS version 20 for windows (SPSS Inc, USA). Malaria parasitaemia and antibody units were log-transformed before analysis in order to obtain approximated Gaussian distributions. Differences in group means were assessed using the one-way analyses of variance (ANOVA) while differences in proportions were assessed using chi-square. Associations between malaria morbidity, anaemia, and antibody levels were assessed by binary logistic regression or multinomial logistic regression. Associations were quantified using odds ratios (OR) with 95% confidence intervals (CI). The levels of correlation between variables were determined by calculating Pearson’s correlation coefficients (r). A difference or correlation giving a P value <0.05 was considered statistically significant.

### Ethical Considerations

This study was conducted as part of a larger project titled “Malaria baseline studies towards characterising and establishing a clinical trial site at Mutenegene, South West Region of Cameroon” between February 2011 and March 2012. Ethical clearance for this study was obtained from the National Ethics Committee in Cameroon (N°198/CNE/SE/2010). Administrative authorisation was obtained from the Ministry of Public Health and the South West Regional Delegation of Public Health. Written informed consent was obtained from the parents or guardians of the children along with the signature of a witness before the children were enrolled into the study. Samples were only collected by trained medical personnel using sterile and pain-reducing equipment.

## Results

### Baseline, six months and twelve months characteristics of the study cohort

Children (374) aged 6 months to 10 years [mean age ± standard error of mean (SEM): 4.39 ± 0.129 years] were enrolled from ten residential quarters in Mutengene. At baseline the proportion of febrile casesm malaria parasitaemic cases and anaemic cases were 20.9%, 18% and 71.4 % (severe: 2.6 %, moderate: 51.9 % and mild: 45.5 %) respectively. After the follow-up (active and passive) of participants for malaria, 32.5 % (116/357) of those who completed the longitudinal study had at least one episode of malaria. The number of malaria attacks ranged from one to four with a mean ± SEM number of 1.44 ± 0.062 malaria attacks per year. The mean ± SEM age of participants who had at least one episode of malaria was 4.06 ± 0.18 years. Malaria parasitaemia positive cases increased from 18 % at enrolment (end of dry season) to 19.3 % 6 months later (rainy season) and then decreased to 12 % 12 months later (dry season). At baseline, 71.5 % of the study participants had anaemia but the number decreased significantly (p < 0.001) to 60.1 % 6 months later and 43.5 % 12 months later (Table 1).

**Table 1.**
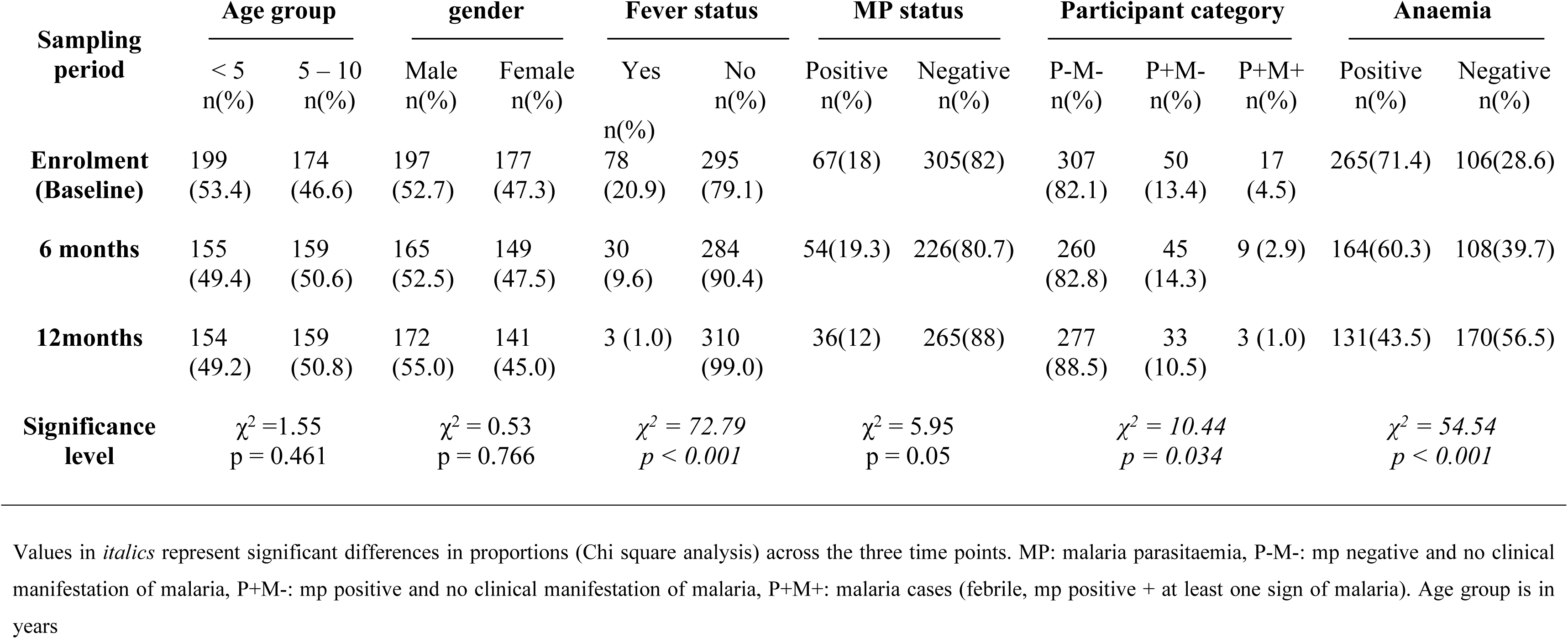
Frequency distribution of participants by age group, gender, fever, malaria parasitaemia status, participant category and anaemic status at rolment, 6 months and 12 months.

### Distribution of antigen-specific antibody responses with age

There was a general increase in total IgG, IgG1, IgG2 and IgG3 antibody levels to all tested antigens with age at all three time points. On a continuous scale, total IgG levels to all four antigens showed a significant positive correlation with age (0.511 ≤ r ≤ 0.261, p < 0.0001) at all three time points (Table 2). Similarly, IgG1 to all the four antigens correlated positively with age at all three time points (0.406 ≤ r ≤ 0.144, p < 0.05) with the exception of GLURP R2 IgG1 which didn’t show any significant correlation with age at 12 months. Also, IgG2 to all the four antigens correlated positively with age at all three time points (0.497 ≤ r ≤ 0213, p < 0.01) with the exception of IgG2 to AMA1-3D7 and GLURP R2 at 12 months. AMA1-3D7 IgG3 did not show any significant correlation with age at any of the three time points. MSP3 IgG3 (r = 0.190, p = 0.014) and GLURP R0 IgG3 (r = 0.275, p < 0.001) levels correlated positively with age only at enrolment while GLURP R2 IgG3 correlated positively with age at all the three time points (0.358 ≤ r ≤ 0.424, p < 0.0001). To further explore the effect of age, antigen – specific antibody responses (mean log values) were compared between the < 5 years and the 5 – 10 years age groups. Mean total IgG antibody levels to all the antigens tested were significantly higher in the 5-10 years age group compared to the < 5 years at all three time points (p < 0.01, ANOVA; Fig 1). At enrolment, IgG1, IgG2 and IgG3 subclasses to all the antigens were also significantly higher (p < 0.05) in the 5-10 years age group compared to the <5 years old with the exception of MSP3 IgG3 (S1 Table). At 6 months GLURP R2 IgG3 in addition to IgG1 and IgG2 subclasses to all the antigens showed a significant difference between the two age groups while at 12 months only AMA1 IgG1, MSP3 IgG2, GLURP R2 IgG3 and GLURP R0 IgG1, 2, & 3 levels were significantly different (p < 0.05). No difference was observed in IgG4 antibodies with age.

**Fig 1.**
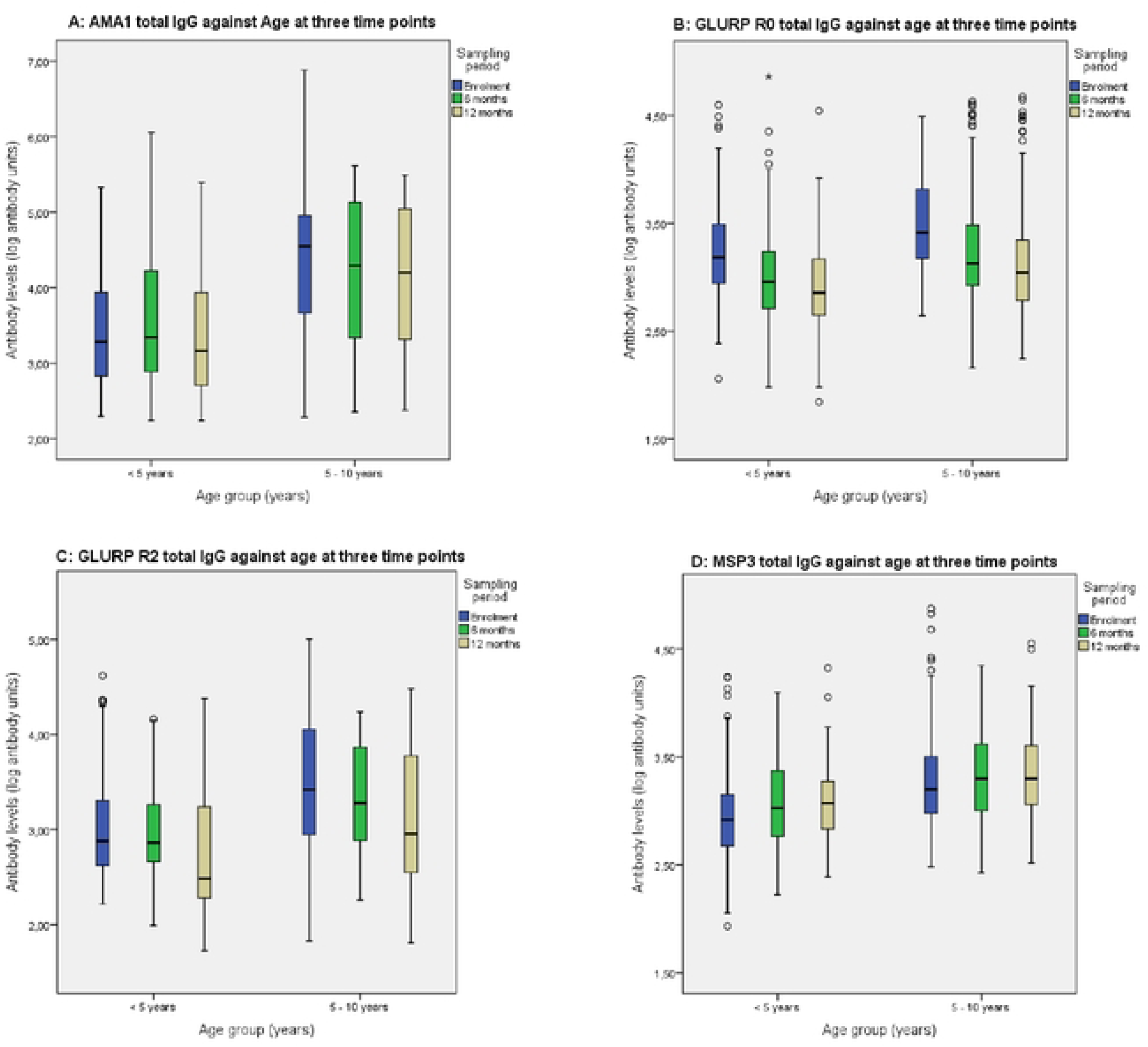
Antigen specific IgG responses with age at baseline, six months and twelve months later. IgG responses to AMA1, GLURP R0, GLURP R2 and MSP3 are shown separately by plots A, B, C and D respectively. IgG antibody units were log transformed and plotted against age groups at enrolment, six months and twelve months

**Table 2.** Correlation of *Plasmodium falciparum* specific total IgG and IgG subclasses with age at three time points in a longitudinal cohort.

| *P. falciparum*<br>Antigens | Total IgG correlation<br>coefficient (n) with age at<br>three time points |  |  | IgG1 correlation coefficient (n)<br>with age at three time points |  |  | IgG2 correlation coefficient (n)<br>with age at three time points |  |  | IgG3 correlation coefficient (n) with<br>age<br>at three time points |  |  |
| --- | --- | --- | --- | --- | --- | --- | --- | --- | --- | --- | --- | --- |
|  | Baselin<br>e | 6<br>months | 12<br>months | Baselin<br>e | 6 months | 12<br>months | Baseline | 6 months | 12<br>months | Baseline | 6<br>months | 12 months |
| AMA1 | 0.511*<br>(348) | 0.450*<br>(257) | 0.399*<br>(250) | 0.406*<br>(247) | 0.344*<br>(157) | 0.270*<br>(179) | 0.438*<br>(308) | 0.277*<br>(159) | NS | NS | NS | NS |
| GLURP R0 | 0.363*<br>(356) | 0.321*<br>(284) | 0.261*<br>(270) | 0.172<sup>‡</sup><br>(354) | 0.203<sup>‡</sup><br>(274) | 0.154<sup>‡</sup><br>(263) | 0.255*<br>(244) | 0.368*<br>(156) | 0.269*<br>(126) | 0.275*<br>(192) | NS | NS |
| GLURP R2 | 0.404*<br>(356) | 0.409*<br>(284) | 0.269*<br>(270) | 0.273*<br>(299) | 0.233*<br>(263) | NS | 0.285*<br>(225) | 0.353*<br>(195) | NS | 0.379*<br>(158) | 0.358*<br>(122) | 0.424* (94) |
| MSP3 | 0.429*<br>(356) | 0.361*<br>(284) | 0.369*<br>(269) | 0.250*<br>(255) | 0.210<sup>‡</sup><br>(230) | 0.144<sup>‡</sup><br>(237) | 0.245<sup>‡</sup><br>(168) | 0.213<sup>‡</sup><br>(227) | 0.497*<br>(265) | 0.190<sup>‡</sup><br>(166) | NS | NS |

Pearson's correlation between antibodies to recombinant *P. falciparum* antigens and age at three time points in a longitudinal cohort of children
\*Correlation significant at $P < 0.001$ , <sup>‡</sup>correlation significant at $P < 0.05$ , NS: correlation not significant. IgG4 to all the antigens showed no correlation with age at any of the three time points and AMA1 IgG3 did not correlate positively with age at any of the three time points.

### IgG, IgG1-4 subclass levels and malaria cases at three time points in the longitudinal cohort

The levels of GLURP R0 IgG (F = 35.7, p <0.001), GLURP R2 IgG (F = 16.5, p <0.001), AMA1-3D7 IgG2 (F = 10.8, p < 0.001) and AMA1-3D7 IgG3 (F = 4.01, p = 0.019) decreased (Fig 2, ANOVA) alongside a significant decrease in the number of febrile (*χ^2^* = 72.8, p < 0.001) and malaria cases (*χ2* = 10.4, p = 0.034) across the three time points (Table 1). This was contrary to the increase observed in total IgG to MSP3 (F = 8.9, p < 0.001) and MSP3 IgG2 levels (F = 44.2, p < 0.001) across the three time points (Fig 2). Parasitaemia prevalence increased from 18 % at baseline to 19.3 % at 6 months and decreased to 12 % at 12 months. Following a similar pattern MSP3 IgG1 (F = 13.9, p < 0.001), MSP3 IgG3 (F = 3.9, p = 0.021), GLURP R0 IgG4 (F = 3.3, p = 0.038) and GLURP R2 IgG2 (F = 9.9, p < 0.001) levels significantly increased from baseline to 6 months and dropped at 12 months. This was contrary to GLURP R0 IgG1 (F = 75.6, p < 0.001) levels which decreased at 6 months and slightly increased at 12 months (Fig 2).

**Fig 2.**
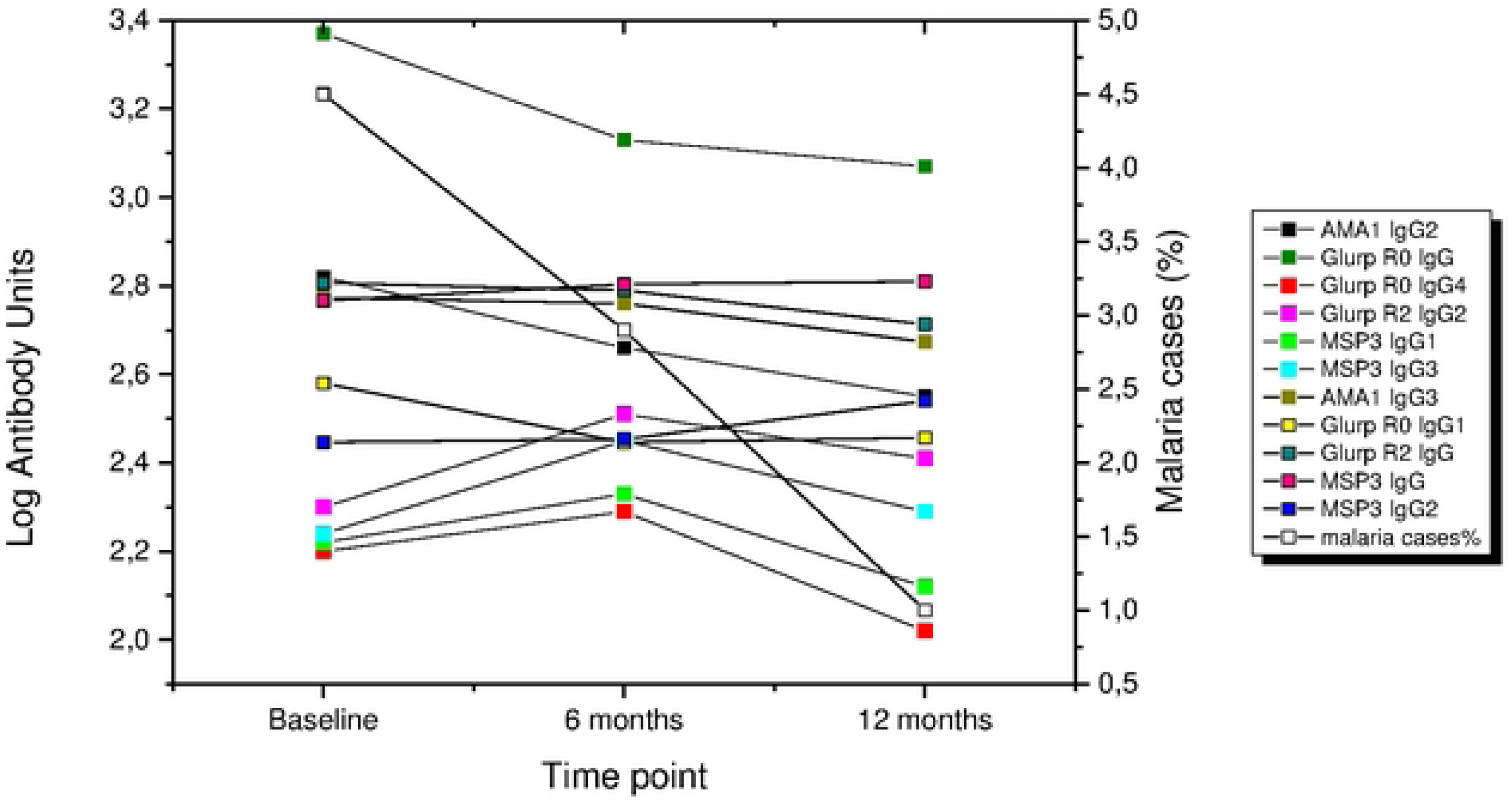
Antibody levels and percentage of malaria cases at three time points in a longitudinal cohort. Antigen specific antibody responses (mean log transformed antibody units) that varied significantly across the three time points (analysis of variance, p<0.05) and a simultaneous decrease in the percentage of malaria cases across the three time points (Chi square analysis, p<0.05). Only antibodies with significant differences are represented here.

### Relationship between antigen specific IgG, IgG1-4 subclasses and *P. falciparum* malaria parasitaemia

Participants were categorized into three groups: those without detectable malaria parasitaemia by microscopy and no clinical manifestation of malaria (P-M-), those who were asymptomatic but were positive for malaria parasitaemia (P+M-) and those who were febrile and malaria parasitaemic plus at least one sign/symptom of malaria (P+M+). Participants who were asymptomatic, but malaria parasitaemia positive (P+M-) were significantly (p < 0.001) older compared to participants in the P+M+ and P-M-groups. In a logistic regression adjusting for age, IgG and all IgG subclasses (except MSP3 IgG2, MSP3 IgG4, AMA1-3D7 IgG4 and GLURP R2 IgG4) to the four antigens tested showed a significant association with protection against malaria parasitaemia at enrolment (Table 3). Generally, cytophilic antibody levels and IgG2 levels (except MSP3 IgG2) were significantly higher (ANOVA) in the P+M-group compared to the P+M+ and P-M-groups at baseline (Fig 3). Using multinomial logistic regression, the children with no parasitaemia (P-M-) were significantly associated with lower antibody levels (total IgG against all four antigens, AMA1-3D7 cytophilic antibodies and GLURP R0 & R2 IgG2 antibodies) compared to children who had malaria episodes (P+M+) at enrolment (Table 4).

**Fig 3.**
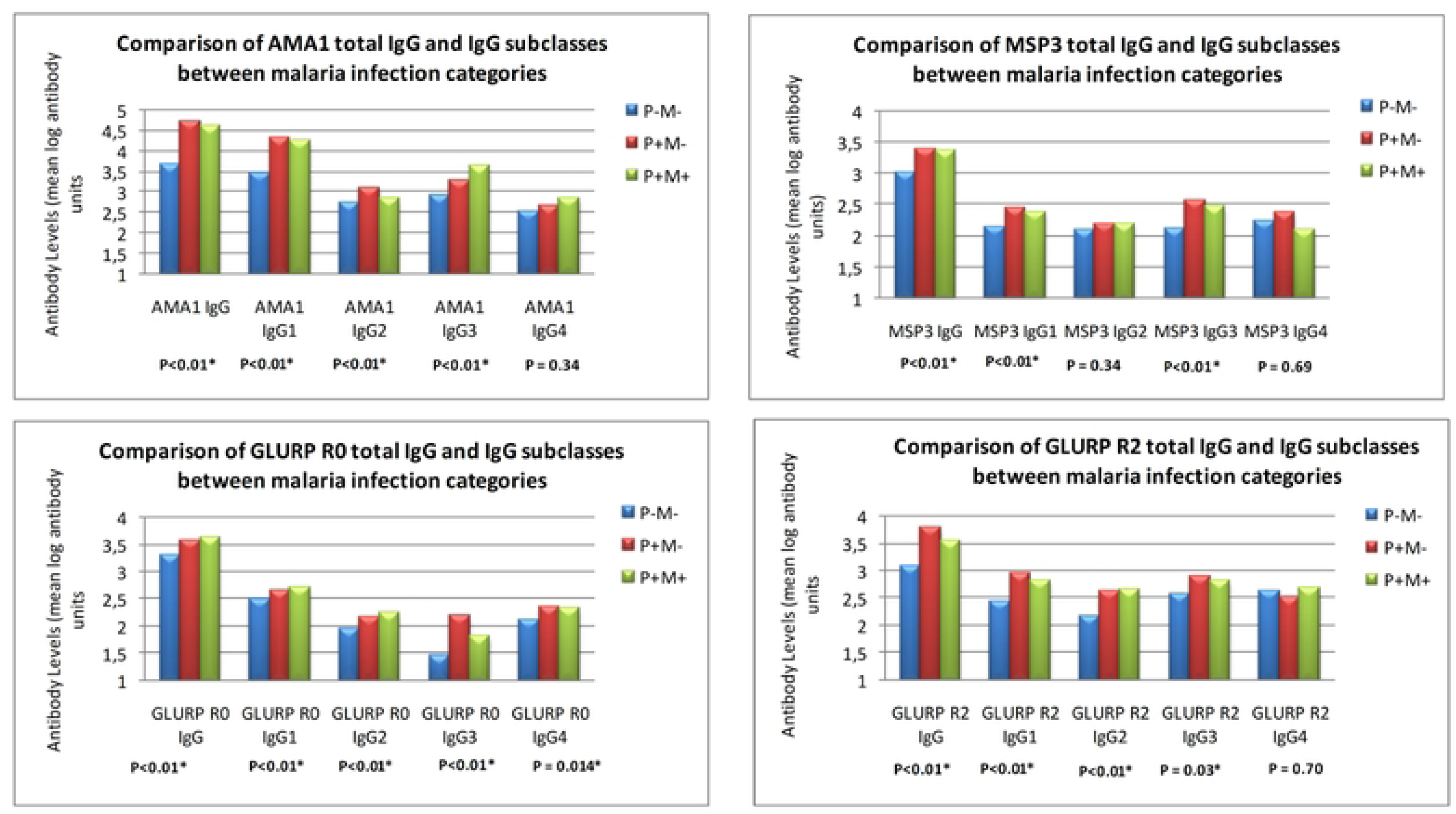
Antibody levels (means) and malaria infection status at enrolment. *statistical significance determined using the one-way analysis of variance, comparing mean values of log transformed antibody units (standard error of means calculated but not indicated here), P+M+:malaria cases (mp positive, febrile plus at least one sign of malaria), P+M-: mp positive but no clinical manifestation of malaria, P-M-: mp negative and no clinical manifestation of malaria

**Table 3.**
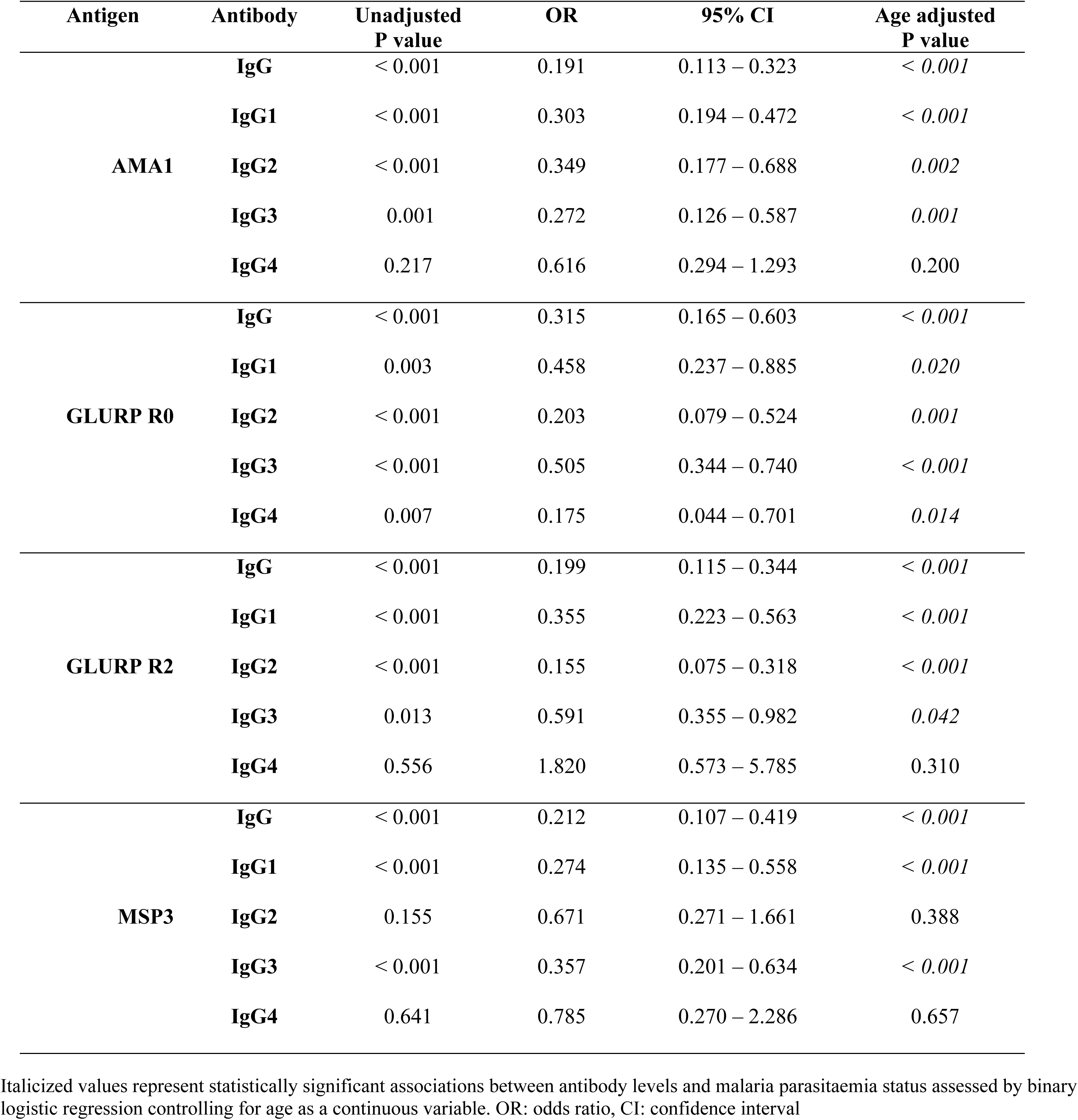
Age adjusted relationship between IgG and IgG subclasses and malaria parasitaemia status at enrolment.

| Antigen | Antibody | Unadjusted<br>P value | OR | 95% CI | Age adjusted<br>P value |
| --- | --- | --- | --- | --- | --- |
| **AMA1** | **IgG** | < 0.001 | 0.191 | 0.113 – 0.323 | < 0.001 |
|  | **IgG1** | < 0.001 | 0.303 | 0.194 – 0.472 | < 0.001 |
|  | **IgG2** | < 0.001 | 0.349 | 0.177 – 0.688 | 0.002 |
|  | **IgG3** | 0.001 | 0.272 | 0.126 – 0.587 | 0.001 |
|  | **IgG4** | 0.217 | 0.616 | 0.294 – 1.293 | 0.200 |
| **GLURP R0** | **IgG** | < 0.001 | 0.315 | 0.165 – 0.603 | < 0.001 |
|  | **IgG1** | 0.003 | 0.458 | 0.237 – 0.885 | 0.020 |
|  | **IgG2** | < 0.001 | 0.203 | 0.079 – 0.524 | 0.001 |
|  | **IgG3** | < 0.001 | 0.505 | 0.344 – 0.740 | < 0.001 |
|  | **IgG4** | 0.007 | 0.175 | 0.044 – 0.701 | 0.014 |
| **GLURP R2** | **IgG** | < 0.001 | 0.199 | 0.115 – 0.344 | < 0.001 |
|  | **IgG1** | < 0.001 | 0.355 | 0.223 – 0.563 | < 0.001 |
|  | **IgG2** | < 0.001 | 0.155 | 0.075 – 0.318 | < 0.001 |
|  | **IgG3** | 0.013 | 0.591 | 0.355 – 0.982 | 0.042 |
|  | **IgG4** | 0.556 | 1.820 | 0.573 – 5.785 | 0.310 |
| **MSP3** | **IgG** | < 0.001 | 0.212 | 0.107 – 0.419 | < 0.001 |
|  | **IgG1** | < 0.001 | 0.274 | 0.135 – 0.558 | < 0.001 |
|  | **IgG2** | 0.155 | 0.671 | 0.271 – 1.661 | 0.388 |
|  | **IgG3** | < 0.001 | 0.357 | 0.201 – 0.634 | < 0.001 |
|  | **IgG4** | 0.641 | 0.785 | 0.270 – 2.286 | 0.657 |

Italicized values represent statistically significant associations between antibody levels and malaria parasitaemia status assessed by binary logistic regression controlling for age as a continuous variable. OR: odds ratio, CI: confidence interval

**Table 4.** Age adjusted relationship between IgG and subclasses antibody levels and malaria clinical status at enrolment.

| Antigen | Antibody | P-M- * |  |  | P+M- * |  |  |
| --- | --- | --- | --- | --- | --- | --- | --- |
|  |  | OR | 95% CI | P value | OR | 95% CI | P value |
| AMA1 | IgG | 0.231 | 0.098 – 0.545 | *0.001* | 1.305 | 0.505 – 3.375 | 0.582 |
|  | IgG1 | 0.341 | 0.155 – 0.79 | *0.007* | 1.17 | 0.485 – 2.826 | 0.727 |
|  | IgG2 | 0.800 | 0.243 – 2.635 | 0.714 | 3.153 | 0.825 – 12.048 | 0.093 |
|  | IgG3 | 0.146 | 0.046 – 0.462 | *0.001* | 0.46 | 0.151 – 1.321 | 0.145 |
|  | IgG4 | 0.434 | 0.127 – 1.479 | 0.182 | 0.648 | 0.189 – 2.223 | 0.490 |
| GLURP<br>R0 | IgG | 0.250 | 0.08 – 0.778 | *0.017* | 0.740 | 0.211 – 2.587 | 0.637 |
|  | IgG1 | 0.371 | 0.119 – 1.155 | 0.087 | 0.755 | 0.214 – 2.667 | 0.663 |
|  | IgG2 | 0.126 | 0.026 – 0.619 | *0.011* | 0.531 | 0.097 – 2.903 | 0.465 |
|  | IgG3 | 0.689 | 0.368 – 1.289 | 0.243 | 1.531 | 0.779 – 3.009 | 0.217 |
|  | IgG4 | 0.188 | 0.017 – 1.437 | 0.176 | 1.009 | 0.096 – 12.559 | 0.940 |
| GLURP<br>R2 | IgG | 0.352 | 0.142 – 0.869 | *0.024* | 2.226 | 0.790 – 6.274 | 0.130 |
|  | IgG1 | 0.454 | 0.199 – 1.038 | 0.061 | 1.383 | 0.568 – 3.369 | 0.476 |
|  | IgG2 | 0.141 | 0.041 – 0.484 | *0.002* | 0.891 | 0.249 – 3.186 | 0.860 |
|  | IgG3 | 0.683 | 0.279 – 1.672 | 0.404 | 1.206 | 0.469 – 3.098 | 0.698 |
|  | IgG4 | 1.00 | 0.470 – 6.787 | 1.00 | 0.469 | 0.063 – 3.492 | 0.460 |
| MSP3 | IgG | 0.217 | 0.072 – 0.655 | *0.007* | 1.046 | 0.327 – 3.340 | 0.940 |
|  | IgG1 | 0.336 | 0.108 – 1.042 | 0.059 | 1.336 | 0.406 – 4.395 | 0.634 |
|  | IgG2 | 0.637 | 0.148 – 2.378 | 0.544 | 0.931 | 0.193 – 4.489 | 0.929 |
|  | IgG3 | 0.408 | 0.162 – 1.023 | 0.056 | 1.187 | 0.469 – 3.003 | 0.717 |
|  | IgG4 | 1.955 | 0.091 – 41.823 | 0.668 | 2.823 | 0.128 – 62.102 | 0.511 |

\*P+M+ is the reference category
Italicized values represent statistically significant comparative associations assessed by multinomial logistic regression using P+M+ as the reference category and controlling for age as a continuous variable. P+M+: malaria cases (mp positive, febrile plus at least one sign of malaria, P+M-: mp positive but no clinical manifestation of malaria, P-M-: mp negative and no clinical manifestation of malaria, OR: odds ratio, CI: confidence interval

### Relationship between antigen specific total IgG, IgG subclasses antibodies and anaemia

At six months, increasing levels of AMA1-3D7 IgG, IgG1 & IgG2, GLURP R0 IgG & IgG1, GLURP R2 IgG, IgG1 & IgG2 and MSP3 IgG, IgG1 & IgG3 were significantly associated with susceptibility to anaemia after controlling for age. However, only AMA1-3D7 total IgG and MSP3 IgG1 remained significant after controlling for age and parasitaemia density (Table 5). This observation was consistent with higher AMA1-3D7 IgG2 (F = 6.26, P = 0.013), GLURP R0 IgG1 (F = 5.41, p = 0.021), GLURP R2 IgG1 (F = 4.865, P = 0.028), GLURP R2 IgG2 (F = 4.417, p = 0.037), MSP3 IgG1 (F = 4.943, p = 0.027) and MSP3 IgG3 (F = 3.989, P = 0.049) mean levels in participants with anaemia compared to those who were not anaemic at six months. At twelve months post enrolment, AMA1-3D7 total IgG (F = 5.368, p = 0.021), IgG1 (F = 5.533, p = 0.020) & IgG3 (F = 5.863, p = 0.019), GLURP R0 IgG3 (F = 5.073, p = 0.027) and MSP 3 IgG1 (F = 8.134, p = 0.005) mean levels were also higher in participants with anaemia compared to those without anaemia (S2 Table). In addition, negative correlation was observed between AMA1-3D7 IgG (r = -0.192, p = 0.002), IgG1 (r = -0.198, p = 0.008) & IgG3 (r = - 0.428, p = 0.001), MSP3 IgG (r = -0.156, p = 0.011) & IgG1 (r = -0.238, p <0.001) and haemoglobin levels at 12 months while GLURP R2 IgG1 (r = -0.145, p = 0.019) correlated negatively with haemoglobin levels at six months. Contrary to this, MSP3 IgG2 (r = 0.136, p = 0.043) levels correlated positively with haemoglobin levels at six months.

**Table 5.**
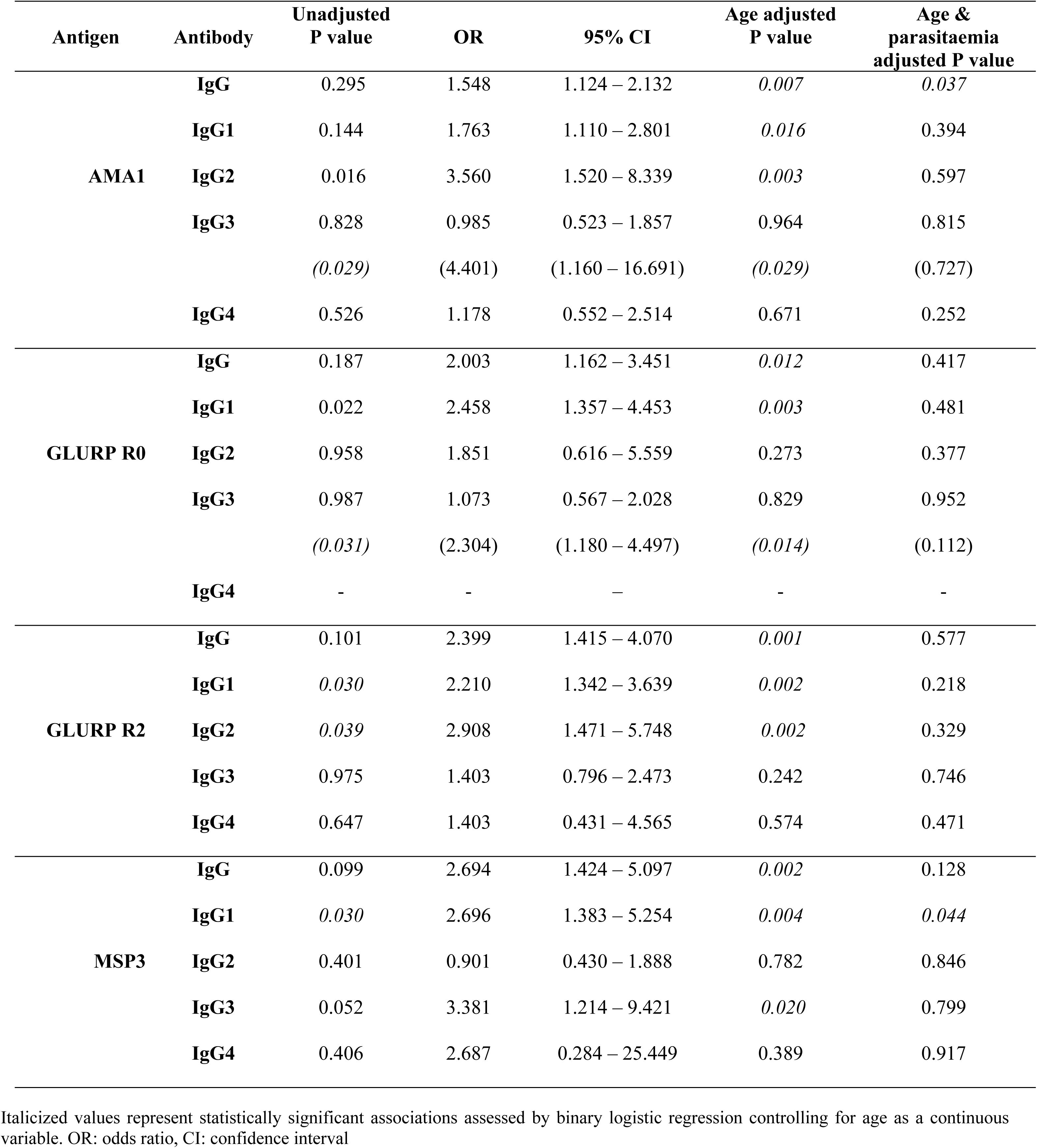
Age and parasitaemia adjusted relationship between IgG and subclasses antibody levels and anaemia status at six months.

## Discussion

This study assessed the levels of total IgG and IgG1-4 subclasses to *P. falciparum* recombinant antigens (AMA1-3D7, GLURP R0, GLURP R2 & MSP3) at three time points in a longitudinal cohort study in relation to malaria morbidity. There was a general increase in total IgG, cytophilic subclasses and IgG2 levels to all the antigens tested with age at all three time points consistent with younger children being more predisposed to malaria episodes compared to older children. Increasing levels of AMA1-3D7 IgG and MSP3 IgG1 were significantly associated with susceptibility to anaemia.

The most predominant IgG subclass antibody type was IgG1 being the highest for all antigens except MSP3 whose IgG3 response was the highest IgG subclass response. This is consistent with results from previous AMA1 [42] and GLURP [43] phase 1 trials which showed that antibodies were predominantly of the IgG1 isotype. Also, high titres of IgG3 antibodies to MSP3 have been shown to be associated with reduced incidence of malaria in longitudinal cohort studies [44].

GLURP R0 IgG, GLURP R2 IgG, AMA1-3D7 IgG2 and AMA1-3D7 IgG3 levels decreased progressively throughout the study accompanied by a corresponding decrease in the proportion of malaria episodes contrary to an increase in MSP3 IgG and MSP3 IgG2 levels across the three sampling time points. The variation of other antibody subclasses (MSP3 IgG1, MSP3 IgG3 AND GLURP R2 IgG2) reflected the increase in malaria parasitaemia prevalence from enrolment (18%) to 19.3% at 6 months and a decrease to 12% at 12 months. *P. falciparum* antibody responses (sporozoites, merozoites, *P. falciparum*-infected erythrocytes & gametocytes) have been shown to rapidly decline after a few months following drug treatment and parasite clearance in study participants with symptomatic infections in both children and adults [7]. MSP3 and GLURP antibodies have been reported to last 6-12 months if no boost infection takes place [45]. A frequent observation in the field is that malaria specific antibody levels are diminished in the absence of parasite exposure and the persistence of antibodies is shorter in children as shown in one study by the fluctuation of antibody levels between seasons [46]. However, the question of longevity of antibody responses is hotly debated with evidence for and against short and long-lived antibody responses [7]. Despite the different patterns of antibody levels observed, it’s evident that the antibody levels are more a reflection of the number of malaria cases and/or malaria parasitaemia density rather than the prevalence. Further clarification on the kinetics and longevity of antibodies could be obtained in a longitudinal study in which the malaria antibody profiles of participants are monitored individually over a defined period.

IgG and IgG subclasses (IgG1, IgG2 & IgG3) to all four recombinant malaria antigens increased with age in accordance with long–standing reports that malaria antibody acquisition is age and exposure dependent. The risk of malaria also decreased with age, as indicated by the fact that the mean age of children who had at least one malaria episode was 4.06 years and children less than 5 years old had more attacks compared to those who were 5–10 years old, as earlier reported on the same cohort [47]. In 2008, a similar study conducted on a cohort of Ghanaian children (3 – 10 years old) showed that isotype and IgG subclasses to AMA1, MSP1_19_, MSP3 and GLURP R0 also increased with age while the risk of malaria decreased with age [41, 48]. Antibody responses against AMA1, MSP2, & MSP3 were also reported to be associated with protection against malaria in a cohort of Kenyan children [49]. In other longitudinal studies in Kenya and Papua New Guinea, older children or children from higher transmission settings had substantially higher antibodies and significant clinical immunity compared with young children [7]. Total IgG, cytophilc subclasses and IgG2 to all four antigens investigated (except MSP3 IgG2) were associated with protection from malaria parasitaemia at enrolment. The levels of these antibodies were higher in children with asymptomatic parasitaemia compared to those with malaria episodes. These observations again agree with previous studies by Dodoo *et al,* [41, 48] which found higher IgG levels to the same antigens to be associated with reduced risk of malaria attacks. In addition, a significant association between GLURP R2 IgG3 and reduced risk of malaria in cohorts of Burkinabe and Ghanaian children was also reported in 2016 [50]. Experimental evidence suggests that AMA1 is involved in merozoite invasion of red blood cells (RBCs) and it is essential to the proliferation and survival of malaria parasites. In a phase I clinical trial with an AMA1 vaccine containing an equal mixture of recombinant proteins from FVO and 3D7 clones of *P. falciparum,* antigen-specific *in vitro* inhibition of both FVO and 3D7 parasites was achieved with IgG purified from sera of vaccinees, demonstrating biological activity of the antibodies [42]. *P. falciparum* GLURP is the target of cytophilic antibodies which are significantly associated with protection against malaria. Immuno-epidemiological studies performed in both high and low malaria transmission areas have revealed a high prevalence of GLURP antibodies in adults as well as an association between high GLURP-specific antibody levels, low parasite densities and protection against malaria [51]. The presence of high titres of IgG3 antibodies against a conserved region of MSP3 has been correlated with protection against the parasite [44, 52]. The promising vaccine candidate GMZ2 which is a fusion protein of *P. falciparum* MSP 3 and GLURP was evaluated in a phase 2b randomized controlled trial in African children and GMZ2 was well tolerated, immunogenic and reduced the incidence of malaria [31].

Some merozoite proteins seem to mediate their protective role through complement-mediated lysis or through cooperation of Fc receptor-bearing cells. In a few instances cytophilic antibodies have been shown to facilitate the phagocytosis of merozoites through opsonization or mediate antibody-dependent cellular inhibition (ADCI). The ADCI effect triggered by merozoite surface components is mediated by the soluble components released by monocytes which inhibit intra-erythrocytic development of the parasite [53]. This mechanism led to the identification of MSP3 and GLURP as major targets of ADCI-effective antibodies [25, 54]. In 2015, Tiendrebeogo and others [55] reported that high ADCI activity was significantly associated with reduced risk against malaria in a longitudinal cohort of Ghanaian children.

Interestingly, increased levels of AMA1-3D7 IgG and MSP3 IgG2 were associated with susceptibility to anaemia. This can be explained by the fact that antibody-mediated immune mechanisms such as phagocytosis of parasitized and unparasitized RBCs have been implicated in the pathogenesis of malarial anaemia [56]. In the process of ridding the system of malaria parasites, the resulting anaemia might be collateral damage. However, MSP3 IgG2 levels correlated positively with haemoglobin levels though no association was observed with regression analysis. It is also worth noting that aside from malaria there were possibly other significant contributors to the high levels of anaemia observed in this study as reported in another paper on the same cohort [47].

## Conclusion

Total IgG, IgG1, IgG3 and IgG2 to all the antigens (except MSP3 IgG2) were associated with protection from malaria parasitaemia and malaria episodes in Cameroonian children. The differing variation in antibody levels to AMA1-3D7, GLURP and MSP3 antigens across the three time points brings to question the longevity of these antibodies with MSP3 IgG apparently persisting longer. Anaemia appears to be the result of collateral damage due the action of malaria parasite specific antibodies but this association warrants further studies.

## Data Availability

All data produced in the present study are available upon reasonable request to the authors

## Acknowledgements

We thank the families from the Mutengene community who consented for their children to participate in this study and the healthcare workers who assisted with the field work. Immense gratitude goes to the Immunology Department of NMIMR, Ghana through its head Professor Ben Gyan for providing the facilities for the antibody assays. We are very grateful towards Professor Michael Theisen of the University of Copenhagen, Denmark for generously providing *P. falciparum* recombinant AMA1-3D7, GLURP R0, GLURP R2 and MSP3 antigens for the antibody ELISAs. Thanks also go to Dr Eric Kyei-Baafour for assisting with the antibody ELISAs and Dr. Bright Adu for his input in drafting this paper.

## Supporting Information

**S1 Table. Total IgG and IgG 1, 2 &3 antibody levels to *P. falciparum* specific antigens compared between the < 5 years and the 5 – 10 years age groups at enrolment, 6 months and 12 months.** Mean log antibody units ± standard error of mean (SEM) compared between the < 5 years and the 5 – 10 years age groups at three time points (enrolment, 6 months, 12 months) by analysis of variance (ANOVA). There was no significant difference in IgG4 (to all four antigens) levels between the two age groups at any of the sampling time points (data not shown here). AMA1: apical membrane antigen 1, GLURP: glutamate rich protein, MSP3: merozoite surface protein 3. *significant difference at P < 0.01, ᶲsignificant difference at P < 0.05

**S2 Table. Mean (±SEM) IgG and IgG1-4 subclass levels and anaemia status at six months and at twelve months.** Comparison of mean ± SEM (standard error of mean) antigen specific antibody levels by anaemia staus at six months and 12 months in some cases; anaemic: haemoglobin < 11g/dl. Values in *italics* represent statistical significance determined using the one-way analysis of variance. *comparison at 12 months that showed a significant difference

## Notes

### Competing Interest Statement

The authors have declared no competing interest.

### Funding Statement

This study was jointly funded by the European and Developing Countries Clinical Trials Partnership (EDCTP) through a Central African Network for Tuberculosis, HIV/AIDS and Malaria (CANTAM) grant awarded to Professor Achidi Eric (EAA) and from an OWSD (Organization for Women in Science for the Developing World) Postgraduate sandwich fellowship awarded to Njua Clarisse Yafi (CN-Y). Research support was also provided in the form of research allowances from Ministry of Higher Education in Cameroon through the Faculty of Science of the University of Yaounde I.

### Author Declarations

Ethical clearance for this study was obtained from the National Ethics Committee in Cameroon (No198/CNE/SE/2010). Administrative authorisation was obtained from the Ministry of Public Health and the South West Regional Delegation of Public Health. Written informed consent was obtained from the parents or guardians of the children along with the signature of a witness before the children were enrolled into the study. Samples were only collected by trained medical personnel using sterile and pain-reducing equipment.

## References

1. Titanji VPK, Tamu VD, Akenji TKN and Joutchop AS. Immunoglobulin G and subclass responses to *Plasmodium falciparum* antigens: A study in highly exposed Cameroonians. Clinical Chemistry and Laboratory Medicine. 2002; 40(9): 937–940.

2. Nkuo-Akenji TK, Ntonifor NN, Ching JK, Kimbi HK, Ndamukong KN, Anong DN, et al. Evaluating a malaria intervention strategy using knowledge, practices and coverage surveys in rural Bolifamba, Southwest Cameroon. Transactions of the Royal Society for Tropical Medicine and Hygiene. 2004; 99:325–332.

3. Wanji S, Kengne-Ouafo AJ, Eyong EEJ, Kimbi HK, Tendongfor N, Ndamukong-Nyanga JL et al. Genetic diversity of *Plasmodium falciparum* merozoite surface protein-1 block 2 sites of contrasting altitudes and malaria endemicities in the Mount Cameroon Region. American Journal of Tropical Medicine and Hygiene. 2012; 86(5): 764–774.

4. Kimbi HK, Sumbele IUN, Nweboh M, Anchang-Kimbi JK, Lum E, Nana Y, et al. Malaria and haematologic parameters of pupils at different altitudes along the slope of mount Cameroon: a cross sectional study. Malaria Journal. 2013; 12:193.

5. Fru-Cho J, Buma VV, Safeukui I, Nkuo-Akenji T, Titanji VPK and Haldar K. Molecular typing reveals substantial *Plasmodium vivax* infection in asymptomatic adults in a rural area of Cameroon. Malaria Journal. 2014; 13:170.

6. WHO. World Malaria Report 2022. 2022a; https://www.who.int/items/global-malaria-programme

7. Fowkes FJI, Boeuf P and Beeson JG. Immunity to malaria in an era of declining malaria transmission. Parasitology. 2016; DOI: 10.1017/S0031182015001249

8. McGregor A and Carrington SP. Treatment of east African *P. falciparum* malaria with west African human gamma-globulin. Transactions of the Royal Society of Tropical Medicine and Hygiene. 1963; 57: 170–175.

9. Sabchareon A, Burnouf T, Ouattara D, Attanath P, Bouharoun-Tayoun H, Chantavanich P et al. Parasitologic and clinical human response to immunoglobulin administration in falciparum malaria. American Journal of Tropical Medicine and Hygiene. 1991; 45: 297–308.

10. Bouharoun-Tayoun H, Oeuvray C, Lunel F and Druilhe P. Mechanisms underlying the monocyte-mediated antibody-dependent killing of Plasmodium falciparum asexual blood stages. Journal of Experimental Medicine. 1995; 182: 409 – 418.

11. de Souza JB, Todd J, Krishegowda G, Gowda DC, Kwiatkowski D and Riley ME. Prevalence and boosting of antibodies to *Plasmodium falciparum* GPIs and evaluation of their association with protection from mild and severe clinical malaria. Infection and Immunity. 2002; 70 (9): 5045–5051.

12. Perlmann P and Bjorkman A. Host parasite interaction and new developments in chemotherapy, immunology and vaccinology. Current Opinion in Infectious Diseases. 2000; 13:431–443.

13. Bereczky S, Montgomery SM, Troye-Blomberg M, Ingegerd R, Shaw M and Farnet A. Elevated anti-malarial IgE in asymptomatic individuals is associated with reduced risk for subsequent clinical malaria. International Journal of Parasitology. 2004; 34:935–942.

14. Ouma C, Keller CC, Opondo DA, Were T, Otieno RO, Otieno MF, et al. Association of Fcγ receptor IIA (CD32) polymorphism with malarial anaemia and high density parasitaemia in infants and young children. Tropical Medicine and Hygiene. 2006; 74(4): 573 – 577.

15. Aucan C, Traore Y, Tall F, Nacro B, Traore-Leroux T, Fumoux F et al. High immunoglobulin IgG(2) and low IgG(4) levels are associated with human resistance to *Plasmodium falciparum* malaria. Infection and Immuinity. 2000; 68:1252–8. doi: 10.1128/IAI.68.3.1252-1258.2000

16. Aucan C, Traoré Y, Fumoux F and Rihet P. Familial Correlation of Immunoglobulin G Subclass Responses to *Plasmodium falciparum* Antigens in Burkina Faso. Infection and Immuninity. 2001; 69(2): 996–1001. doi: 10.1128/IAI.69.2.996-1001.2001

17. WHO. Position Paper: Malaria Vaccine; Weekly Epidemiological Record No 9, 97: 61 – 80. 2022b; http://www.who.Int/wer. Accessed 24 August, 2023

18. WHO. RTS,S Malaria Vaccine, News Room Q/A. 2023; https://www.who.int/news-room/questions&answers/item/q-a-on-rts-s-malaria-vaccine. Accessed 24 August, 2023

19. Tamborrini M, Schafer A, Hauser J, Zou L, Paris D and Pluschke G. The malaria blood stage antigen PfCyRPA formulated with the TLR-4 agonist adjuvant GLA-SE elicits parasite growth inhibitory antibodies in experimental animals. Malaria Journal. 2023; 22:210. 10.1186/s12936-023-04638-8

20. Crompton DP, Pierce KS and Miller LH. Advances and challenges in malaria vaccine development. The Journal of Clinical Investigation. 2010a; 120(12): 4168–4178.

21. Hill VSA. Vaccines against malaria. Philosophical Transactions of the Royal Society B. 2011; 366: 2806–2814.

22. Sirima SB, Nébié I, Ouédraogo A, Tiono AB, Konaté AT, Gansané A et al. Safety and immunogenicity of the *Plasmodium falciparum* merozoite surface protein-3 long synthetic peptide (MSP3-LSP) malaria vaccine in healthy, semi-immune adult males in Burkina Faso, West Africa. Vaccine. 2007; 25(14):2723–32

23. Sirima SB, Tiono AB, Ouédraogo A, Diarra A, Ouédraogo AL, Diarra A et al. Safety and Immunogenicity of the Malaria Vaccine Candidate MSP3 Long Synthetic Peptide in 12–24 Months-Old Burkinabe Children. PLoS ONE. 2009; 4(10): e7549. doi:10.1371/journal.pone.000754

24. Koram KA, Adu B, Ocran J, Karikari YS, Adu-Amankwah S, Ntiri M, et al. Safety and Immunogenicity of EBA-175 RII-NG Malaria Vaccine Administered Intramuscularly in Semi-Immune Adults: A Phase 1, Double-Blinded Placebo Controlled Dosage Escalation Study. PLoS ONE. 2016; 11 (9): e0163066. doi:10.1371/journal.pone.0163066

25. Theisen M, Soe S, Oeuvray C, Thomas AW, Vuust J, Danielsen S et al. The glutamate-rich protein (GLURP) of Plasmodium falciparum is a target for antibody-dependent monocyte-mediated inhibition of parasite growth in vitro. Infection and Immunity. 1998; 66: 11–17.

26. Sheehy SH, Duncan CJ, Elias SC, Collins KA, Ewer KJ, Spencer AJ et al. Phase Ia clinical evaluation of the *Plasmodium falciparum* blood-stage antigen MSP1 in ChAd63 and MVA vaccine vectors. The American Society of Gene and Cell Therapy. 2011; (12):2269–76. doi: 10.1038/mt.2011.176.

27. Sheehy SH, Duncan CJA, Elias SC, Biswas S, Collins KA, O’Hara GA et al. Phase Ia Clinical Evaluation of the Safety and Immunogenicity of the *Plasmodium falciparum* Blood-Stage Antigen AMA1 in ChAd63 and MVA Vaccine Vectors. PLoS ONE. 2012; 7(2): e31208. doi:10.1371/journal.pone.0031208

28. Chitnis CE, Mukherjee P, Mehta S, Yazdani SS, Dhawan S, Shakri AR et al. Phase I Clinical Trial of a Recombinant Blood Stage Vaccine Candidate for Plasmodium falciparum Malaria Based on MSP1 and EBA175. PLoS ONE. 2015; 10(4): e0117820. doi:10.1371/journal.pone.0117820

29. Ogutu BR., Apollo OJ, McKinney D, Okoth W, Siangla J, Dubovsky F, et al. Blood Stage Malaria Vaccine Eliciting High Antigen-Specific Antibody Concentrations Confers No Protection to Young Children in Western Kenya. PLoS ONE. 2009; 4(3): e4708. doi:10.1371/journal.pone.0004708

30. Sagara I, Dicko A, Ellis RD, Fay MP, Diawara SI, Assadou MH et al. A randomized controlled phase 2 trial of the blood stage AMA1-C1/Alhydrogel malaria vaccine in children in Mali. Vaccine. 2009; 27(23): 3090–3098. doi:10.1016/j.vaccine.2009.03.014.

31. Sirima SB, Mordmüller B, Milligan P, Ngoa UA, Kironde F, Atuguba F et al A phase 2b randomized, controlled trial of the efficacy of the GMZ2 malaria vaccine in African children. Vaccine. 2016; 34 (38):4536–4542. doi: 10.1016/j.vaccine.2016.07.041

32. Salamanca DR, Gómez M, Camargo A, Cuy-Chaparro L, Molina-Franky J, Reyes C et al. *Plasmodium falciparum* Blood Stage Antimalarial Vaccines: An Analysis of Ongoing Clinical Trials and New Perspectives Related to Synthetic Vaccines. Frontiers in Microbiology. 2019; 10:2712. doi: 10.3389/fmicb.2019.02712

33. El-Moamly AA and El-Sweify MA. Malaria vaccines: the 60-year journey of hope and final success – lessons learned and future prospects. Tropical Medicine and Health. 2023;51:29. 10.1186/s41182-023-00516-w

34. Shi YP, Udhayakumar V, Oloo AJ, Nahlen BL and Lal AA. Differential effect and interaction of monocytes, hyperimmune sera,and immunoglobulin G on the growth of asexual stage *Plasmodium falciparum* parasites. American Journal of Tropical Medicine and Hygiene. 1999; 60: 135–141.

35. Wanji S, Tanke T, Atanga SN, Ajonina C, Nicholas T and Fontenille D. *Anophheles* species of the Mount Cameroon Region; biting habits, feeding behaviour and entomological inoculation rates. Tropical Medicine and International Health. 2003; 8:643–9.

36. Cameroon Development Corporation (CDC). Meteorological service, Esuke Station, 2011. http://www.cdc-cameroon.com/subpages. Accessed 20 June 2014.

37. Tanga MC, Ngundu WI, Tchouassi PD. Daily survival and human blood index of major malaria vectors associated with oil palm cultivation in Cameroon and their role in malaria transmission. Tropical Medicine & International Health. 2011; 16:447–57.

38. Bigoga JD, Manga L, Titanji VPK, Coetzee M and Leke RGF. Malaria vectors and transmission dynamics in coastal South-west Cameroon. Malaria Journal. 2007; 6:5

39. WHO. Haemoglobin concentrations for the diagnosis of anaemia and assessment of severity. Vitamin and Mineral Nutrition Information System. World Health Organization, Geneva. 2011; http://www.who.int/vmins/indicators/haemoglobin. Accessed 24 May 2013.

40. Cheesbrough M. District Laboratory Practice in Tropical Countries, Cambridge:Part 2. Cambridge University Press, England. 2000; pp338–340.

41. Dodoo D, Aitkins A, Kusi KA, Lamptey H, Remarque E, Milligan P et al. Cohort study of the association of antibody levels to AMA1, MSP1_19_, MSP3 and GLURP with protection from clinical malaria in Ghanaian children. Malaria Journal. 2008; 7: 142.

42. Malkin EM, Diemert DJ, McArthur JH, Perreault JR., Miles AP, Giersing BK, et al. Phase 1 clinical trial of Apical Membrane Antigen 1: an asexual blood-stage vaccine for *Plasmodium falciparum* malaria. Infection and Immunity. 2005; 73(6): 3677–3685.

43. Hermsen CC, Verhage DF, Telgt DSC, Teelan K, Bousema TJ, Roestenberg M, et al. Glutamate-rich protein (GLURP) induces antibodies that inhibit in vito growth of *Plasmodium falciparum* in a phase 1 malaria vaccine trial. Vaccine. 2007; 25(15): 2930–2940.

44. Polley DS, Tetteh KAK, Lloyd MJ, Akpogheneta JO, Greenwood MB, Bojang AK et al. *Plasmodium falciparum* merozoite surface protein 3 is a target of allele-specific immunity and alleles are maintained by natural selection. Journal of Infectious Diseases. 2007; 195: 279–287.

45. Bauman A, Magda MM, Urbaez M, Vivas-Martinez S, Duran R, Tahidid N et al. Naturally acquired immune responses to malaria vaccine candidate antigens MSP3 and GLURP in Guahibo and Piaroa indigenous communities of the Venezuelan Amazon. Malaria Journal. 2012; 11: 46.

46. Achtman AH, Bull PC, Stephens R and Langhorne J. Longevity of the immune response and memory to blood-stage malaria infection. CTMI. 2005; 297: 71–102.

47. Njua-Yafi C, Achidi EA, Anchang-Kimbi JK, Apinjoh TO, Mugri RN, Chi HF, et al. Malaria, helminths, co-infection and anaemia in a cohort of children from Mutengene, South Western Cameroon. Malaria Journal. 2016; 15:69. DOI: 10.1186/s12936-016-1111-2

48. Dodoo D, Atuguba F, Bosomprah S, Ansah NA, Lamptey H, Egyir B et al. Antibody levels to multiple malaria vaccine candidate antigens in relation to clinical malaria episodes in children in the Kasena-Nankana District of Northern Ghana. Malaria Journal. 2011; 10: 108.

49. Osier FH, Mackinnon MJ, Crosnier C, Fegan G, Kamugu G, Wanaguru M, et al. New antigens for multi-component blood-stage vaccines against *Plasmodium falciparum* malaria. Sci Transl Med. 2014; 6(247): 247ra102. Doi:10.1126/scitranslmed.3008705

50. Adu B, Cherif MK, Bosomprah S, Diarra A, Arthur FKN, Dickson EK, et al. Antibody levels against GLURP R2, MSP1 block 2 hybrid and AS202.11 and the risk of malaria in children living in hyperendemic (Burkina Faso) and hypoendermic (Ghana) areas. Malaria Journal. 2016; 15:123. DOI: 10.1186/s12936-016-1146-4

51. Pratt-Riccio LR, Perce-da-Silva D, Josué da Costa L, Theissen M, Santos F, Tadeu DC et al. Genetic polymorphisms in the glutamate-rich protein of *Plasmodium falciparum* field isolates from a malaria-endemic area of Brazil. Mem Inst Oswaldo Cruz. 2013; 108(4): 523–528.

52. Mazumdar S, Mukherjee P, Yazdani SS, Jain SK, Mohammed A and Virander SC. *Plasmodium falciparum* merozoite surface protein 1 (msp-1) – msp-3 chimeric protein: Immunogenicity determined with human-compartible adjuvants and induction of protective immune response. Infection and Immunity. 2010; 78(2): 872–883.

53. Imam M, Yengkhom SD, Akhilesh KV and Virander SC. Comparative immunogenicities of full-length *Plasmodium falciparum* merozoite surface protein 3 and a 24-kilodalton N-terminal fragment. Clinical and Vaccine Immunology. 2011; 18(8): 1221–1228.

54. Oeuvray C, Bouharoun-Tayoun H, Gras-Masse H, Boyyius E, Kaidoh T, Aikawa M, et al. Merozoite surface protein-3: a malaria protein inducing antibodies that promote *Plasmodium falciparum* killing by coorperation with blood monocytes. BLOOD. 1994; 84(5): 1594–1602.

55. Tiendrebeogo RW, Adu B, Singh SK, Dziegiel MH, Nebie I, Christiansen M et al. Antibody-dependent cellular inhibition is associated with reduced risk against febrile malaria in a longitudinal cohort study involving Ghanaian children. Brief Report, Open Forum Infectious Diseases. 2015; DOI:10.1093/ofid/ofv044

56. Nussenblatt V, Musaka G, Metzger A, Ndeezi G, Garrett E and Semba R. Anaemia and IL-10, TNF-α and erythropoietin levels among children with acute, uncomplicated *Plasmodium falciparum* malaria. Clinical and Diagnostic Laboratory Immunology. 2001; 8(6): 1164–1170.

